# Estimating Incidence of Respiratory Syncytial Virus- and Influenza Virus-Associated Hospitalizations with Community-Acquired Pneumonia and Other Acute Respiratory Infection Among Adults in Japan During and After the COVID-19 Era: A Multicenter Active Surveillance Study (APSG-J2)

**DOI:** 10.1101/2025.06.18.25329683

**Authors:** Haruka Maeda, Shingo Masuda, Bhim Gopal Dhoubhadel, Yuka Fujita, Yuji Akiba, Yutaka Nishigaki, Kei Nakashima, Hiroyuki Ito, Masayuki Nogi, Yoshihito Otsuka, Masayuki Ishida, Eiji Takeuchi, Norichika Asoh, Toyomitsu Sawai, Koichi Hayakawa, Eileen M Dunne, Claudia Schwarz, Bradford D. Gessner, Elizabeth Begier, Shuhei Ito, Ataru Igarashi, Shinobu Osanai, Konosuke Morimoto, Koya Ariyoshi, Adult Pneumonia Study Group-Japan 2 (APSG-J2)

**Author notes:** **Corresponding author** Konosuke Morimoto, Address: Department of Respiratory Infections, Institute of Tropical Medicine, Nagasaki University, 1-12-4 Sakamoto, Nagasaki, 852-8523, Japan. **Main point** This multicenter study estimated RSV and influenza-associated hospitalization incidences among adults aged ≥25 years in Japan (2022–2024). Among those aged ≥65 years, RSV-associated CAP/ARI incidences were 29 and 36 per 100,000 person-years in 2022–2023 and 2023–2024, respectively.

## Abstract

**Background:** Quantifying the burden of respiratory syncytial virus (RSV) in adults is challenging compared to influenza, and data among older adults remain scarce in Japan. Country-specific evidence is essential to support RSV vaccination policy.

**Methods:** This prospective, multicenter study (APSG-J2) targeted hospitalized adults with community-acquired pneumonia (CAP) and other acute respiratory infection (ARI) in seven community-hospitals across four catchment areas in Japan between September 2022 and August 2024. Respiratory samples were analyzed using a multiplex polymerase chain reaction (PCR) kit to detect RSV and influenza. Incidence rates of RSV- and influenza-associated hospitalizations were estimated using study data and national statistics, stratified by age and region.

**Results:** Among 3,047 hospitalized patients with CAP/ARI, 1,502 (49.3%) underwent multiplex PCR testing. RSV and influenza were detected in 2.8% and 3.3% of tested patients, respectively. The incidences of RSV-associated CAP/ARI hospitalizations among adults aged ≥65 years were 29 and 36 per 100,000 person-years in the first and second years, respectively, with higher incidences among those aged ≥85 years (150 and 131 per 100,000 person-years). Influenza incidence increased markedly in the second year (from 11 to 71 per 100,000 person-years for adults age ≥65 years), possibly reflecting post-COVID-19 transmission changes.

**Conclusions:** This is the first active surveillance study in Japan to estimate RSV- and influenza-associated hospitalization incidence among adults during and after the COVID-19 era. The results may still be influenced by the lingering effects of COVID-19 restrictions on social interaction. Continued surveillance is essential to accurately assess RSV burden in the adult population.

## 1. Introduction

Acute respiratory infections (ARIs) are a leading cause of morbidity and mortality, particularly among older adults (1). Respiratory syncytial virus (RSV) is a well-recognized viral pathogen primarily known for affecting infants (2, 3). Recent evidence suggests a significant burden among older adults (4–6). However, quantifying the true burden of RSV in adults is challenging compared to influenza, as RSV testing is not routinely performed in clinical practice, and testing sensitivity in adults is lower compared to infants due to the reduced viral load and others factors (7, 8). Much of the previous evidence has relied on modeling methods using time-series analysis (9–13), and retrospective studies based on clinical diagnoses (14–16). Ideal studies require active surveillance using laboratory diagnostic tests to avoid underestimating the true burden of RSV infection (17–20).

To date, in Japan, two kinds of RSV vaccines were approved for use in adults aged ≥60 years (21). Neither of the RSV vaccines for older adults has yet been incorporated into Japan’s National Immunization Program (NIP), while influenza vaccines for this age group have been included in the NIP since 2001. Country-specific evidence on disease burden is essential to support public health decision-making regarding RSV vaccine policy for older adults. Additionally, baseline data on disease burden prior to vaccine implementation are crucial for evaluating the potential impact of vaccine introduction. However, data on burden of RSV among older adults in Japan remain scarce, especially regarding RSV-associated hospitalizations (22–25).

Our research team previously conducted the Adult Pneumonia Study Group–Japan (APSG-J) surveillance between 2012 and 2014 at four hospitals located on each of Japan’s major islands (26). This multicenter study contributed critical insights into the epidemiology of the community-acquired pneumonia (CAP) in adults. During this study period, RSV was detected in 4.3% and influenza A/B in 4.7% among 2,037 adults aged ≥65 years with CAP (27). Since that initial surveillance, multiple changes, including the COVID-19 pandemic, have influenced the epidemiological landscape of respiratory infections in Japan. In light of these changes, we initiated the second phase of the APSG-J surveillance (APSG-J2), aiming to reassess the incidence and etiological spectrum of CAP and other ARI.

In this study, we aimed to estimate the incidence of RSV- and influenza-associated CAP and other ARI hospitalizations based on the APSG-J 2 surveillance conducted between September 1, 2022, and August 31, 2024. We also assessed the clinical characteristics and outcomes of affected patients.

## 2. Methods

### 2.1. APSG-J2 Surveillance

The APSG-J2 is an ongoing, prospective, multicenter, hospital-based study launched in September 2022 in adults in Japan. It is being conducted in seven community hospitals located in four catchment areas, known as ‘Niji-Iryouken’ (secondary medical areas): Asahikawa, Kamogawa, Kochi, and Nagasaki. These areas, selected from Japan’s four main islands, have populations ranging from approximately 30,000 to 400,000, except Kamogawa, with a population of <30,000. The study targets hospitalized adults aged ≥18 years diagnosed with CAP and other ARI. Detailed inclusion and exclusion criteria are provided in Supplementary Section 1.

Supplementary Figure 1 outlines the screening and enrollment process conducted by study staff and clinicians at each site. For patients with whom consent could not be coordinated, enrollment followed an “opt-out” approach in line with Japanese research ethics guidelines. For individuals who declined participation after being informed, minimal data (age, sex, and diagnosis) were collected to support incidence estimation. Demographic and clinical data were collected from electronic medical records and interviews with participants or proxies and recorded in an electronic case report form using REDCap (28).

This study includes patients enrolled between September 1, 2022, and August 31, 2024, a period that partially overlapped with strict public health measures implemented under Japan’s Infection Control Law in response to the COVID-19 pandemic. These measures remained in place until May 8, 2023, when the government reclassified COVID-19 as a Category V infectious disease, equivalent to seasonal influenza.

### 2.2. Multiplex PCR for Respiratory Bacteria and Virus

For patients who provided informed consent for specimen collection, respiratory samples, such as sputum, saliva, or nasopharyngeal swabs, were collected and analyzed using a commercially available multiplex polymerase chain reaction (PCR) respiratory panel (Fast-Track Diagnostics® Respiratory Pathogen 33 assay, Siemens Healthineers, Germany) at a commercial laboratory (LSI Medience, Japan). This test detects 33 kinds of respiratory pathogens, including RSV (subtypes A and B) and influenza A and B.

### 2.3. Definitions

A case of CAP was defined as an ARI with new pulmonary infiltrates on chest X-ray or CT consistent with pneumonia, including increased pulmonary density, alveolar infiltrates with air bronchograms, or pleural effusion. Patients without present radiological evidence of CAP were categorized as other ARI. Hospitalized cases of CAP and other ARIs were considered RSV- or influenza-positive if any collected specimens tested positive for RSV, influenza A, or B by the multiplex PCR.

### 2.4. Statistical analysis

Continuous variables are presented as medians with interquartile ranges (IQRs), while categorical variables are as counts and proportions. Demographics and characteristics of hospitalized patients with CAP and other ARIs who underwent multiplex PCR and those tested positive for RSV or influenza are described.

Incidence rates were calculated separately for two enrollment periods: September 2022–August 2023 and September 2023–August 2024. To estimate the incidence of hospitalized CAP and other ARI as well as those associated with RSV and influenza, we utilized study data alongside publicly available national statistics, applying a modified model used in a previous study (26). The total number of hospitalized CAP and other ARI hospitalizations, both overall, by study sites at each catchment area, and by study year, was estimated by combining data from three arms (Supplementary Figure 1): (1) patients who consented, (2) patients included in the surveillance arm under opt-out consent, and (3) patients who declined but had minimal data collection. The total number of CAP and other ARI hospitalizations by age group for each study year was obtained from study sites in each catchment area (Figure 1A). We then calculated the proportion of hospitalized CAP and other ARI among all hospitalizations for each year at study sites in each catchment area by dividing the number of CAP and other ARI hospitalizations by the total number of hospitalizations at the same study sites in each catchment area during the corresponding year. To account for patients residing outside the designated catchment area, we adjusted the denominator by multiplying the total number of hospitalizations by one minus the proportion of patients from outside the catchment area (29). For all catchment areas except Kamogawa (which had only one participating hospital), data from two hospitals were aggregated to calculate these proportions. These proportions were then multiplied by the total number of hospitalizations in each respective catchment area, based on government statistics, to estimate the total number of CAP and other ARI hospitalizations per year for each catchment area. Catchment area hospitalization data were derived from the Patient Survey conducted by the Ministry of Health, Labour and Welfare between September 1 and 30, 2023 (29). As the survey captured the number of discharged patients for the month of September only, this was used as a proxy for hospitalization figures. To estimate annual hospitalizations in each catchment area, monthly hospitalization numbers were estimated by applying a ratio of September hospitalizations to those in the corresponding month at the national-level, and then summed the adjusted monthly numbers (30). Since monthly hospitalization data were available only at the national level, this ratio was uniformly applied across all catchment areas. To focus on general medical beds, we excluded discharges from psychiatric beds, long-term care beds, and tuberculosis-designated beds by adjusting the total using the respective proportions (29). The final estimate of annual CAP and other ARI hospitalizations was then divided by the population of the corresponding catchment areas based on 2020 national census data to calculate incidence rates (31, 32). These estimates were stratified by age group to provide age-specific incidence. Population data for catchment areas were provided in 5-year age intervals. As only data for individuals aged ≥25 years were available, incidence estimates were restricted to this population. Incidence was reported for the following age groups: ≥25, ≥65, ≥75, 65–74, 75–84, and ≥85 years. Annual incidence rates are expressed per 100,000 population, with corresponding 95% confidence intervals (CIs).

**Figure 1.**
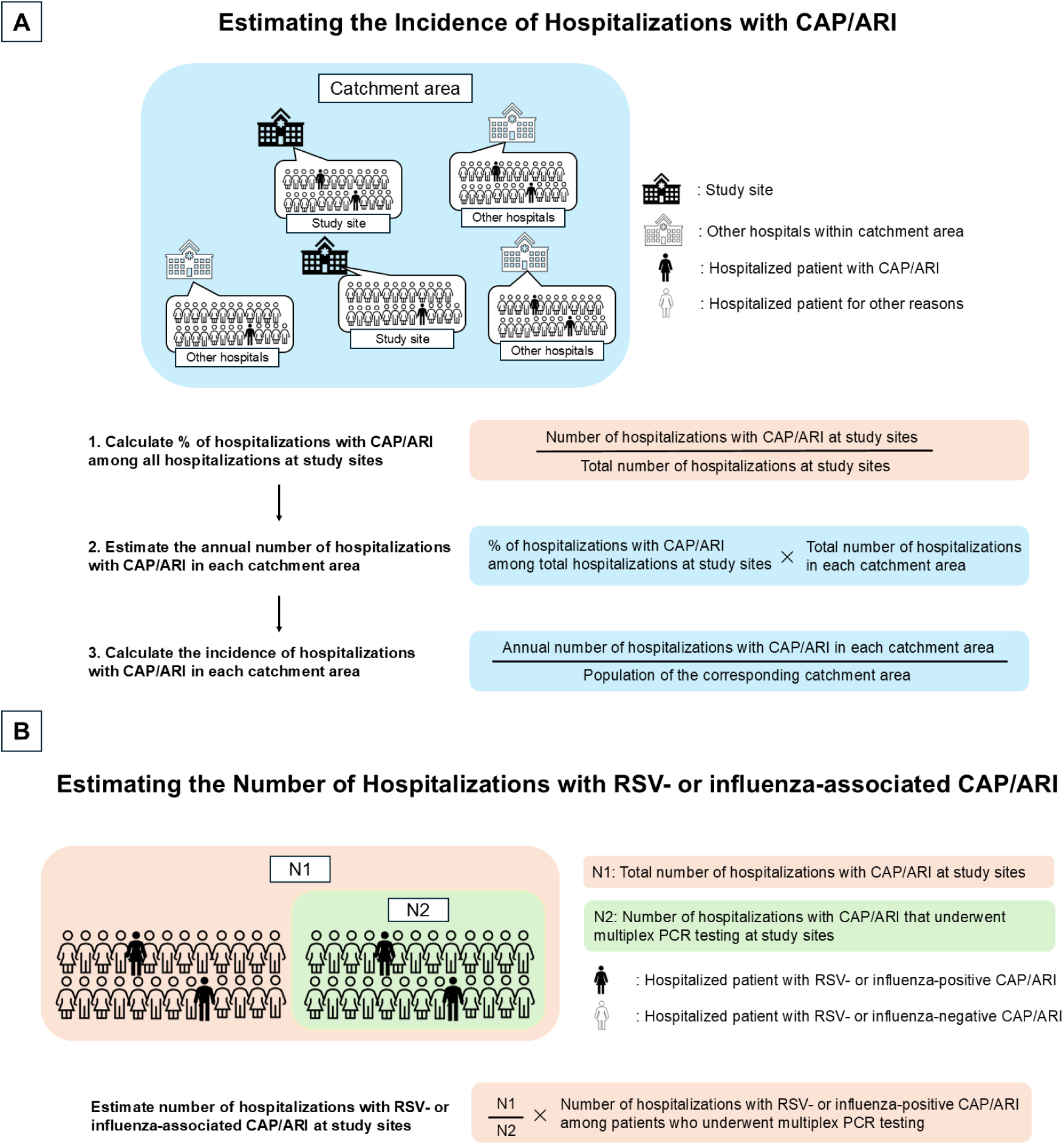
(A) Overview of the steps to estimate the incidence of hospitalizations with CAP and other ARI in each catchment area. First, the percentage of hospitalizations with CAP and other ARI at study sites within each catchment area was calculated by dividing the number of hospitalizations with CAP and other ARI at study sites by total number of hospitalizations at the corresponding sites. This percentage was then applied to the entire catchment area by multiplying it by the total number of hospitalizations in each catchment area to estimate the annual number of hospitalizations with CAP and other ARI in each catchment area. Finally, the incidence of hospitalizations with CAP and other ARI in each catchment area was calculated by dividing the estimated annual number of hospitalizations with CAP and other ARI in each catchment area by the population of the corresponding catchment area. (B) Overview of estimating the number of hospitalizations with RSV- or influenza-associated CAP and other ARI at study sites. Abbreviations: CAP, community-acquired pneumonia; ARI, acute respiratory infection.

To estimate incidence rates of RSV- or influenza-associated CAP and other ARI hospitalizations, the same method described above was applied, except that the total number of RSV- or influenza-associated hospitalizations were used in place of the total number of CAP and other ARI hospitalizations. To calculate the number of RSV- or influenza-associated CAP and other ARI hospitalizations by age group for each study year by study sites in each catchment area, we multiplied the number of RSV- or influenza-positive CAP and other ARI hospitalizations among those who underwent multiplex PCR by the ratio of the total number of CAP and other ARI hospitalizations to the number of such hospitalizations with multiplex PCR (Figure 1B). This approach assumes that patients who were not tested had similar characteristics (and therefore a similar proportion of RSV or influenza positivity) to those who underwent testing.

All analyses were conducted using Stata version 17.0 (StataCorp, College Station, TX, USA).

### 2.5. Ethics

This study was approved by the Institutional Review Board of the Institute of Tropical Medicine, Nagasaki University with a centralized review (Approval No.: 220616247).

## 3. Results

A total of 3,047 hospitalizations with CAP and other ARI were identified across four catchment areas between September 1, 2022, and August 31, 2024: 1,408 in the first year and 1,639 in the second year. Multiplex PCR results were available for 1,502 patients (49.3%) (Figure 2). The characteristics of patients who underwent testing were generally similar to those who did not (Supplementary Section 2). Among the 1,502 tested patients, sputum, saliva, and nasopharyngeal swab were collected from 1,328 (88.4%), 386 (25.7%), and 108 (7.2%) patients, respectively (Supplementary Table 2).

**Figure 2.**
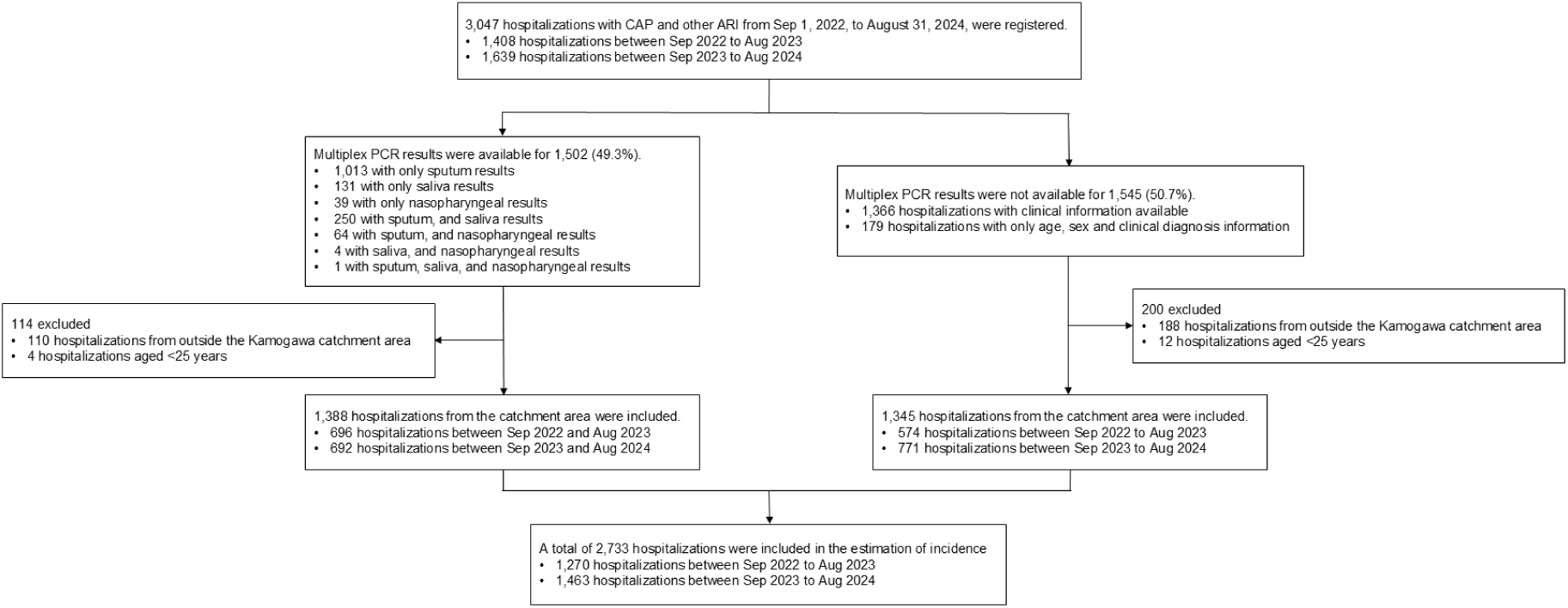
Flowchart of patient inclusion for CAP and other ARI from September 1, 2022, to August 31, 2024, in Japan. A total of 3,047 hospitalizations were registered, of which multiplex PCR results were available for 1,502 (49.3%) hospitalizations. Among all registered hospitalizations, 2,733 hospitalizations were in the analysis for incidence estimation. Abbreviations: CAP, community-acquired pneumonia; ARI, acute respiratory infection.

### 3.1 Characteristics of patients hospitalized with CAP and other ARI who underwent multiplex PCR

Table 1 shows the characteristics of 1,502 patients hospitalized with CAP and other ARI who underwent multiplex PCR. The median age was 81 years (IQR: 73–88), and 64.1% were male. Overall, 82.2% had at least one underlying condition, 17.4% were nursing home residents, and 36.5% required some form of care (Supplementary Table 3). The most common underlying conditions were chronic heart disease (40.4%), chronic lung disease (40.2%), and diabetes mellitus (23.2%). Patient distribution was similar across the two years. Clinically, 7.9% were admitted to ICU, 6.5% required mechanical ventilation, and in-hospital mortality was 8.0%. The median length of hospitalization was 15 days (IQR: 10–27), and 90.6% were diagnosed with CAP (Table 1).

**Table 1.**
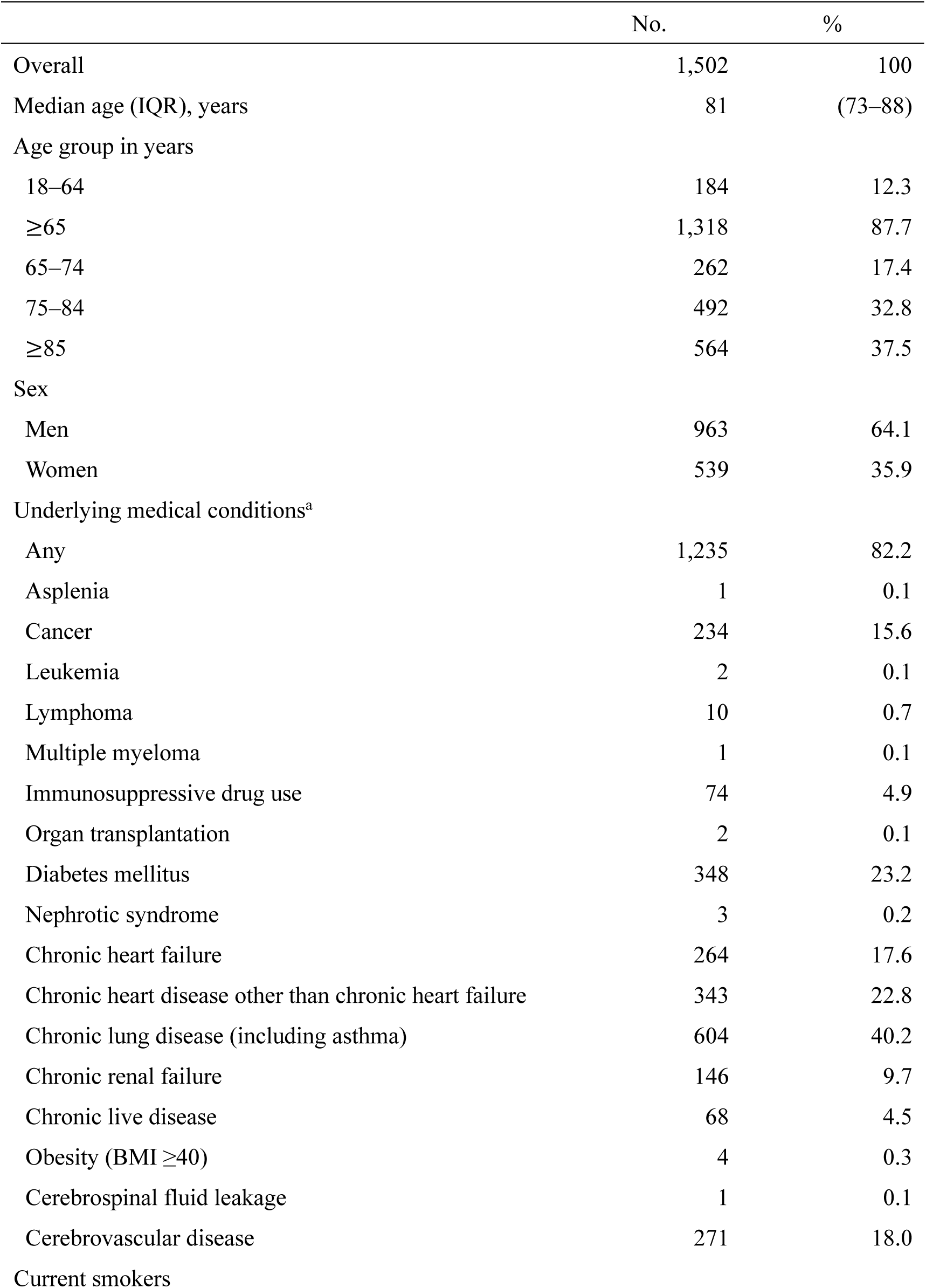
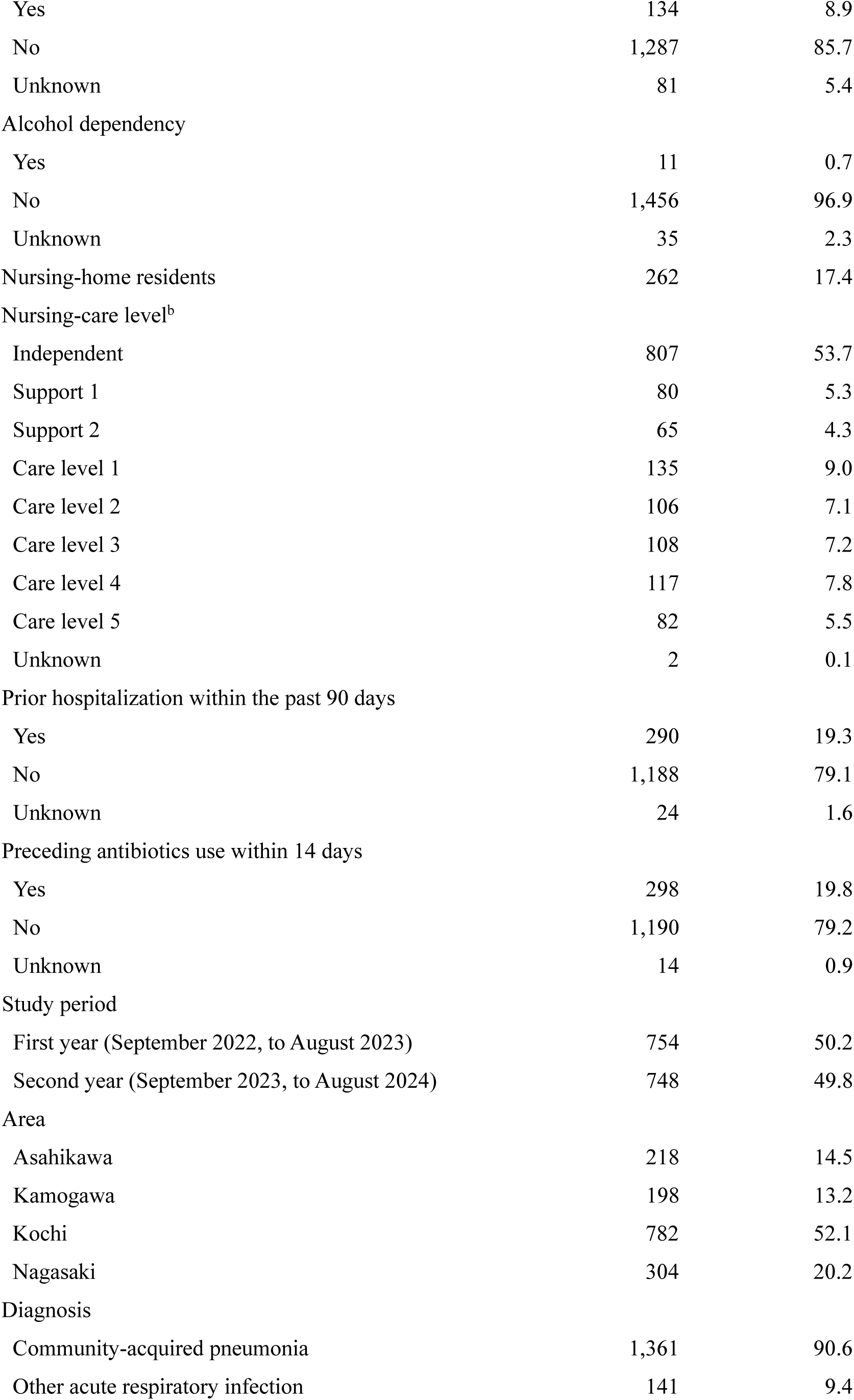
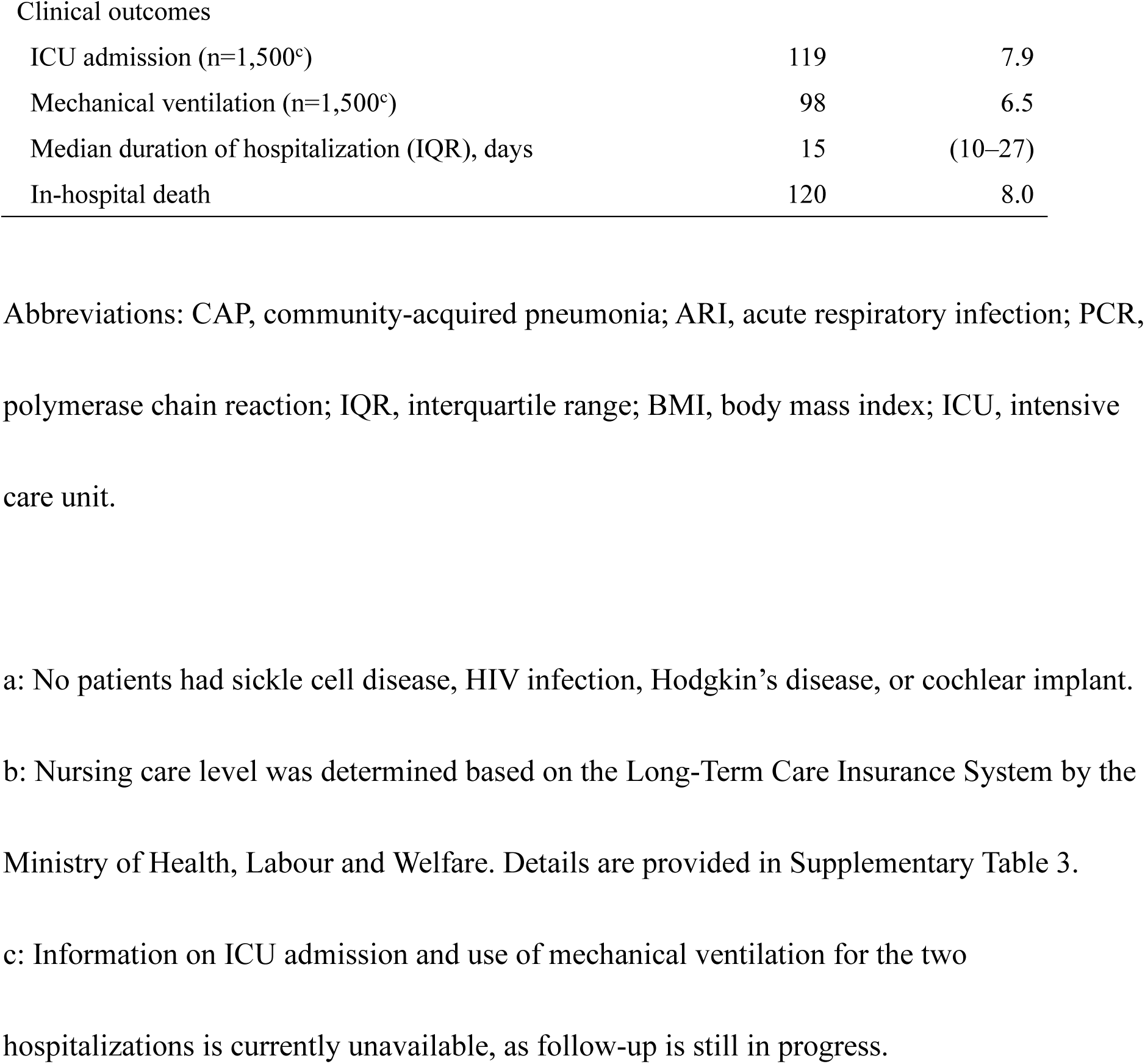
Characteristics of adults hospitalized with CAP and other ARI who underwent multiplex PCR in Japan, September 2022–August 2024.

|  | No. | % |
| --- | --- | --- |
| Overall | 1,502 | 100 |
| Median age (IQR), years | 81 | (73–88) |
| Age group in years |  |  |
| 18–64 | 184 | 12.3 |
| ≥65 | 1,318 | 87.7 |
| 65–74 | 262 | 17.4 |
| 75–84 | 492 | 32.8 |
| ≥85 | 564 | 37.5 |
| Sex |  |  |
| Men | 963 | 64.1 |
| Women | 539 | 35.9 |
| Underlying medical conditions<sup>a</sup> |  |  |
| Any | 1,235 | 82.2 |
| Asplenia | 1 | 0.1 |
| Cancer | 234 | 15.6 |
| Leukemia | 2 | 0.1 |
| Lymphoma | 10 | 0.7 |
| Multiple myeloma | 1 | 0.1 |
| Immunosuppressive drug use | 74 | 4.9 |
| Organ transplantation | 2 | 0.1 |
| Diabetes mellitus | 348 | 23.2 |
| Nephrotic syndrome | 3 | 0.2 |
| Chronic heart failure | 264 | 17.6 |
| Chronic heart disease other than chronic heart failure | 343 | 22.8 |
| Chronic lung disease (including asthma) | 604 | 40.2 |
| Chronic renal failure | 146 | 9.7 |
| Chronic liver disease | 68 | 4.5 |
| Obesity (BMI ≥40) | 4 | 0.3 |
| Cerebrospinal fluid leakage | 1 | 0.1 |
| Cerebrovascular disease | 271 | 18.0 |
| Current smokers |  |  |
| Yes | 134 | 8.9 |
| No | 1,287 | 85.7 |
| Unknown | 81 | 5.4 |
| Alcohol dependency |  |  |
| Yes | 11 | 0.7 |
| No | 1,456 | 96.9 |
| Unknown | 35 | 2.3 |
| Nursing-home residents | 262 | 17.4 |
| Nursing-care level<sup>b</sup> |  |  |
| Independent | 807 | 53.7 |
| Support 1 | 80 | 5.3 |
| Support 2 | 65 | 4.3 |
| Care level 1 | 135 | 9.0 |
| Care level 2 | 106 | 7.1 |
| Care level 3 | 108 | 7.2 |
| Care level 4 | 117 | 7.8 |
| Care level 5 | 82 | 5.5 |
| Unknown | 2 | 0.1 |
| Prior hospitalization within the past 90 days |  |  |
| Yes | 290 | 19.3 |
| No | 1,188 | 79.1 |
| Unknown | 24 | 1.6 |
| Preceding antibiotics use within 14 days |  |  |
| Yes | 298 | 19.8 |
| No | 1,190 | 79.2 |
| Unknown | 14 | 0.9 |
| Study period |  |  |
| First year (September 2022, to August 2023) | 754 | 50.2 |
| Second year (September 2023, to August 2024) | 748 | 49.8 |
| Area |  |  |
| Asahikawa | 218 | 14.5 |
| Kamogawa | 198 | 13.2 |
| Kochi | 782 | 52.1 |
| Nagasaki | 304 | 20.2 |
| Diagnosis |  |  |
| Community-acquired pneumonia | 1,361 | 90.6 |
| Other acute respiratory infection | 141 | 9.4 |
| Clinical outcomes |  |  |
| ICU admission (n=1,500<sup>c</sup>) | 119 | 7.9 |
| Mechanical ventilation (n=1,500<sup>c</sup>) | 98 | 6.5 |
| Median duration of hospitalization (IQR), days | 15 | (10–27) |
| In-hospital death | 120 | 8.0 |

Abbreviations: CAP, community-acquired pneumonia; ARI, acute respiratory infection; PCR, polymerase chain reaction; IQR, interquartile range; BMI, body mass index; ICU, intensive care unit.
a: No patients had sickle cell disease, HIV infection, Hodgkin's disease, or cochlear implant.
c: Information on ICU admission and use of mechanical ventilation for the two hospitalizations is currently unavailable, as follow-up is still in progress.

### 3.2. Patients hospitalized with CAP and other ARI positive for RSV and influenza

Figure 3 shows the monthly percentage of RSV- and influenza-positive hospitalizations among CAP and other ARI patients who underwent multiplex PCR. Overall, 42 of 1,502 patients (2.8%) tested positive for RSV: 2.3% and 3.3 % in the first and second year, respectively. Influenza was detected in 49 patients (3.3%): 0.8% and 5.7% in the first and second year, respectively. Two distinct peaks in influenza-positive hospitalizations were observed: a larger peak during the 2023–2024 winter and a smaller peak in March 2023. In contrast, RSV-positive hospitalizations exhibited multiple, less distinct peaks without a clear seasonal pattern with 10% proportion of positive cases in the highest month.

**Figure 3.**
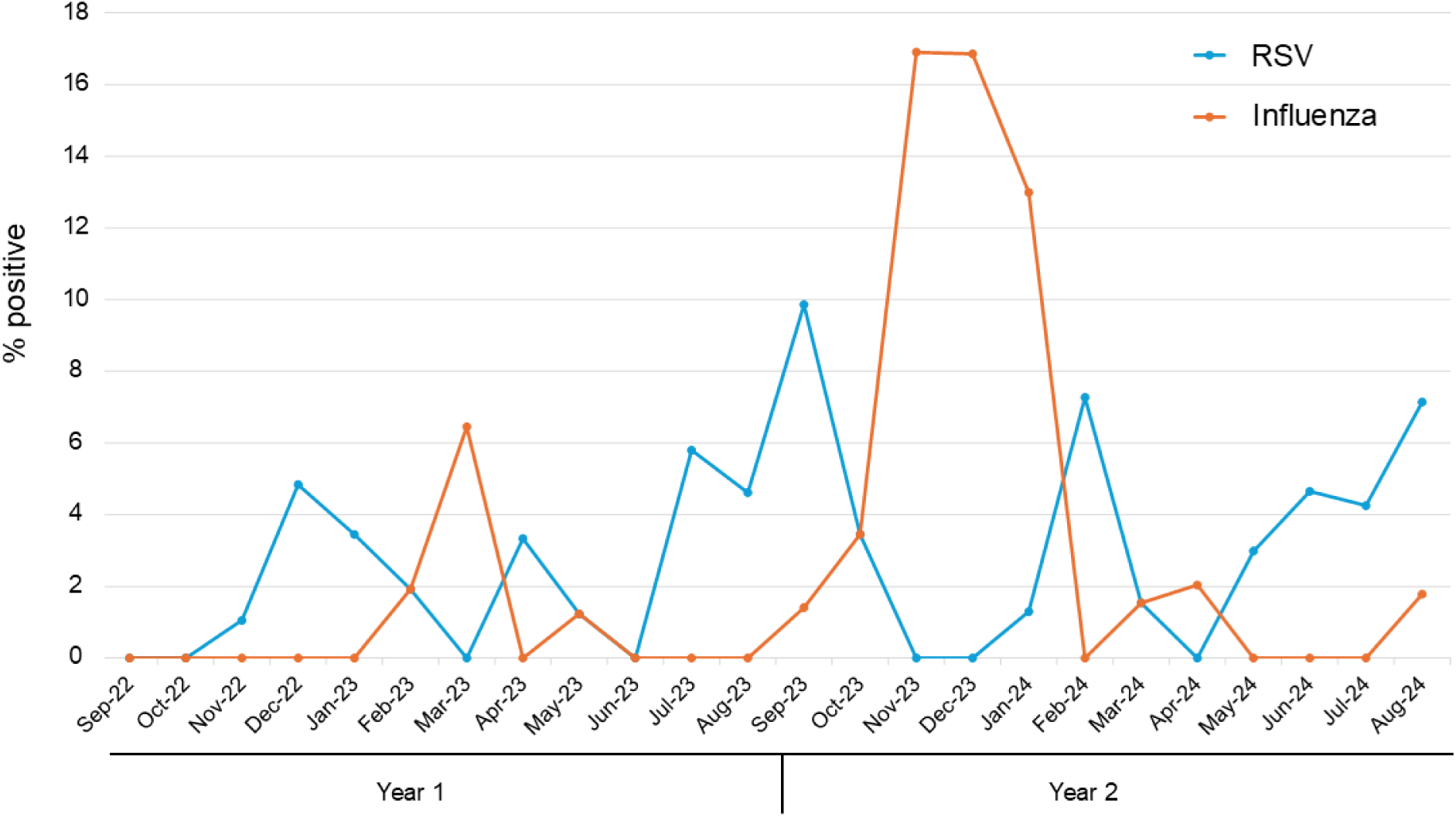
Monthly percentage of RSV- and influenza-positive hospitalizations among community-acquired pneumonia and other acute respiratory patients who underwent multiplex PCR from September 2022 to August 2024 in Japan. The blue line represents the percentage of RSV-positive hospitalizations, while the orange line represents influenza-positive hospitalizations. Abbreviations: RSV, respiratory syncytial virus.

Table 2 summarizes characteristics of RSV- and influenza-positive hospitalizations. Compared with influenza-positive patients, those with RSV were older, more likely to be female, and more frequently residents of nursing home. CAP diagnoses were more frequent among RSV-positive patients. Clinically, ICU admission was required in two RSV-positive patients (4.8%), whereas none among influenza-positive patients. Mechanical ventilation was administered to two patients in each group (4.8% for RSV, 4.1% for influenza). In-hospital deaths occurred in three RSV-positive (7.1%) and one influenza-positive patient (2.0%).

**Table 2.**
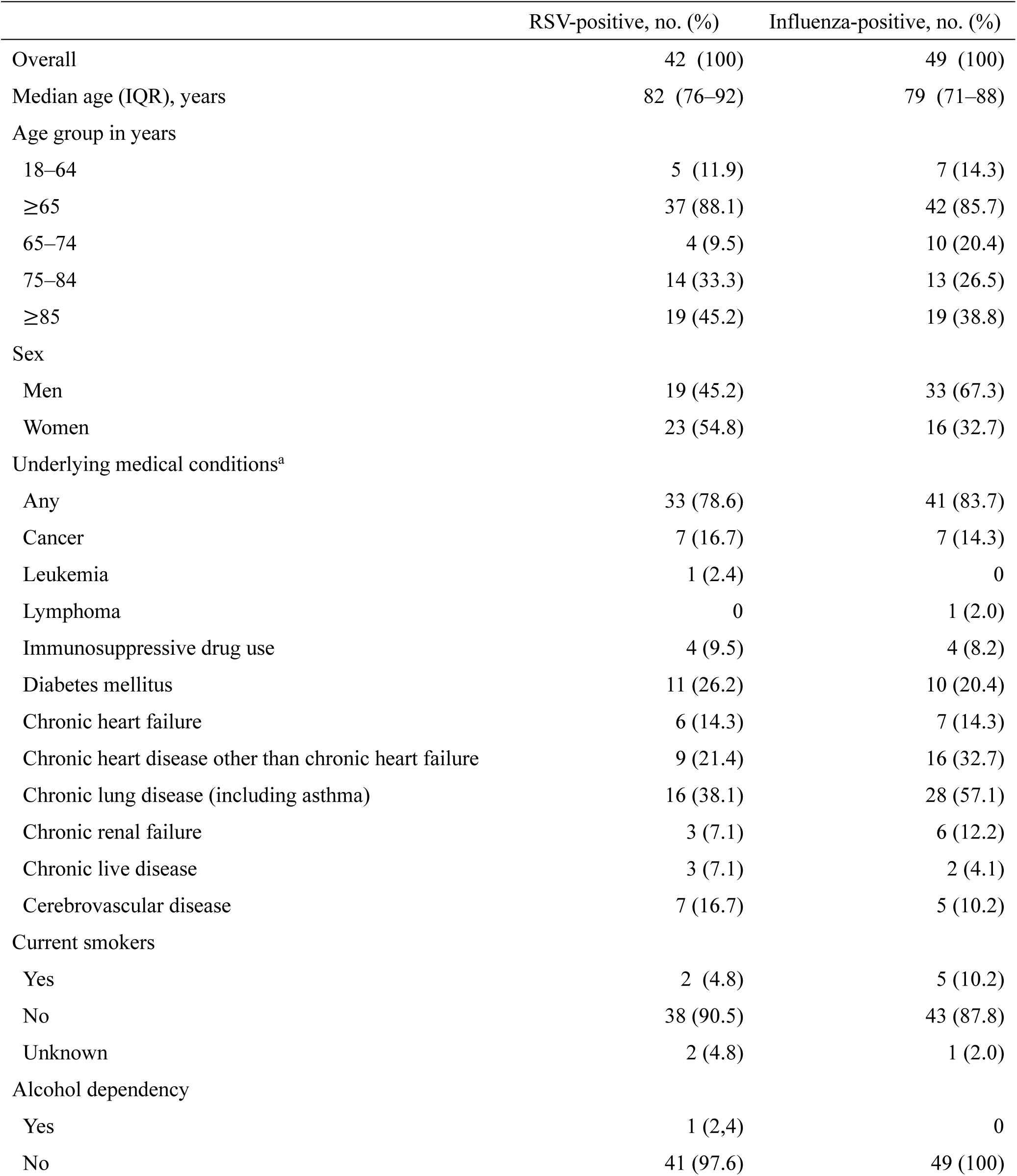
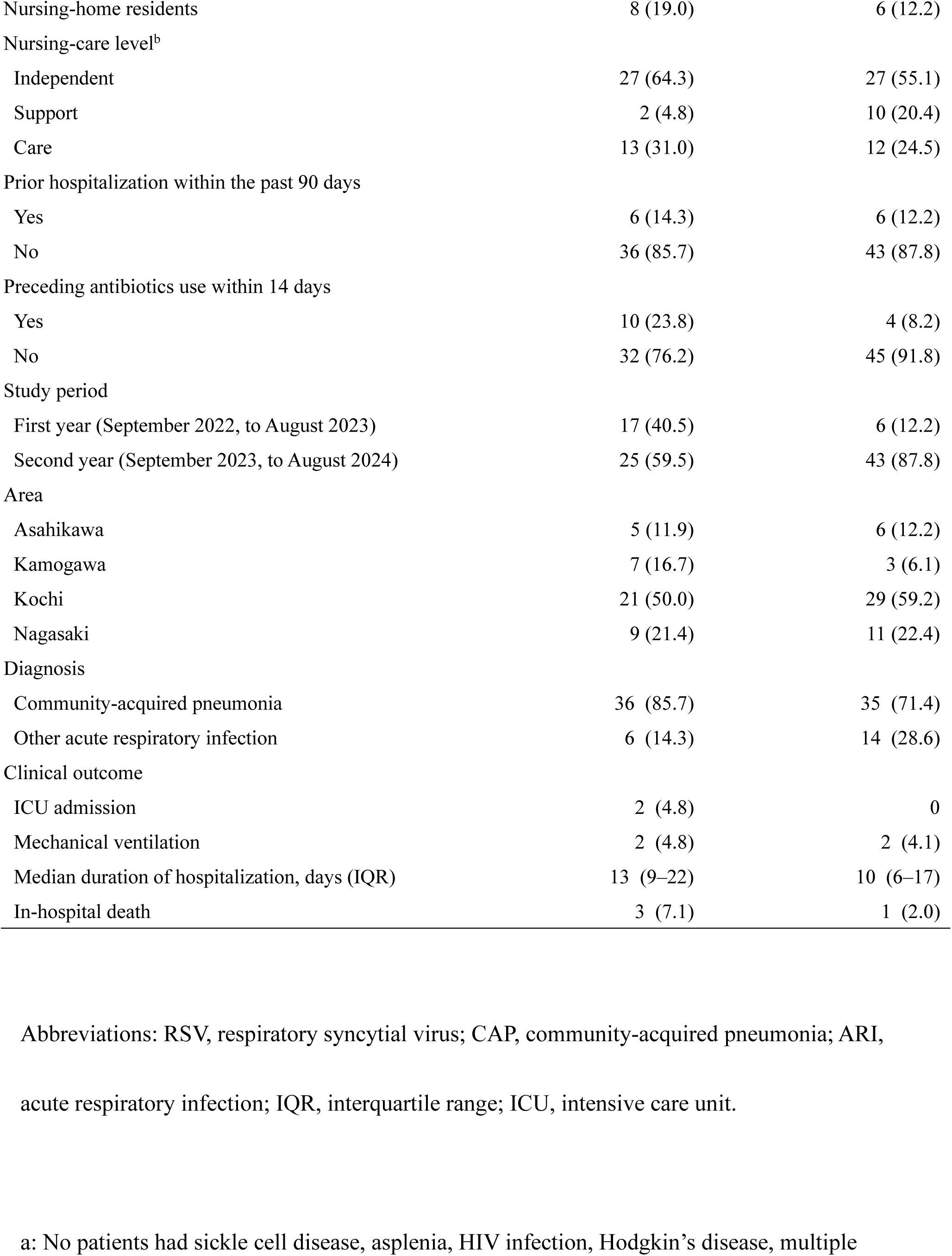
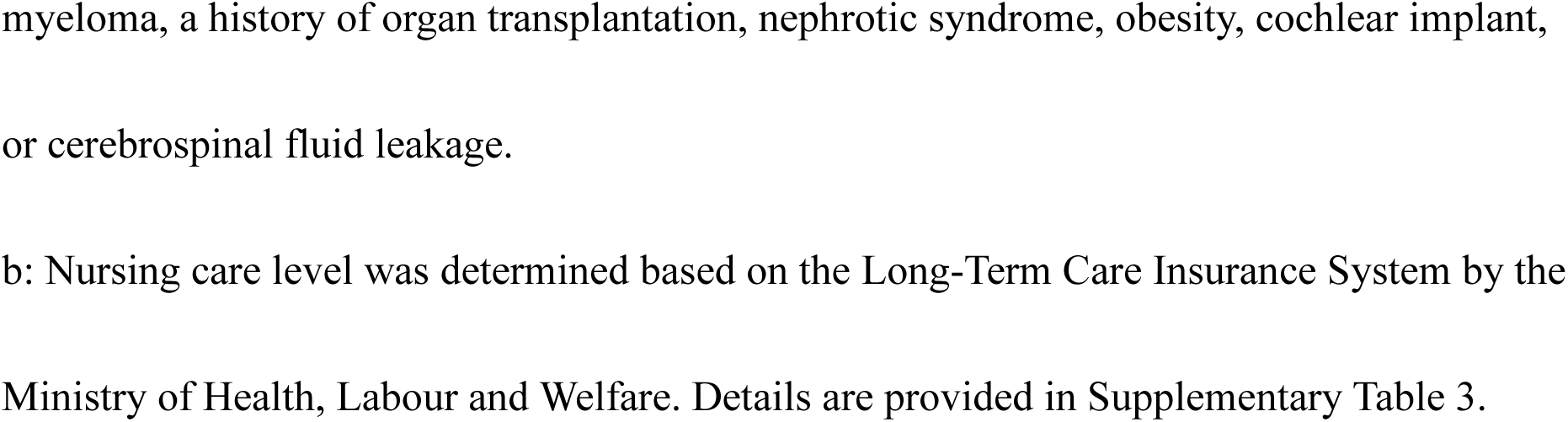
Characteristics of adults hospitalized with RSV- and influenza-positive CAP and other ARI in Japan, September 2022–August 2024.

### 3.3. Incidence of total, RSV-associated, and influenza-associated CAP and other ARI

Table 3 presents the incidence rates of hospitalizations for total CAP and other ARI, as well as those associated with RSV or influenza. Among adults aged ≥25 years, the incidence of total CAP and other ARI hospitalizations per 100,000 person-years was 483 (95% CI: 456– 509) and 539 (95% CI: 511–566) per 100,000 person-years in the first and second year, respectively. Among adults aged ≥65 years, corresponding incidence rates were 1,113 (95% CI: 1,050–1,176) and 1,217 (95% CI: 1,152–1,281) per 100,000 person-years, respectively.

**Table 3.**
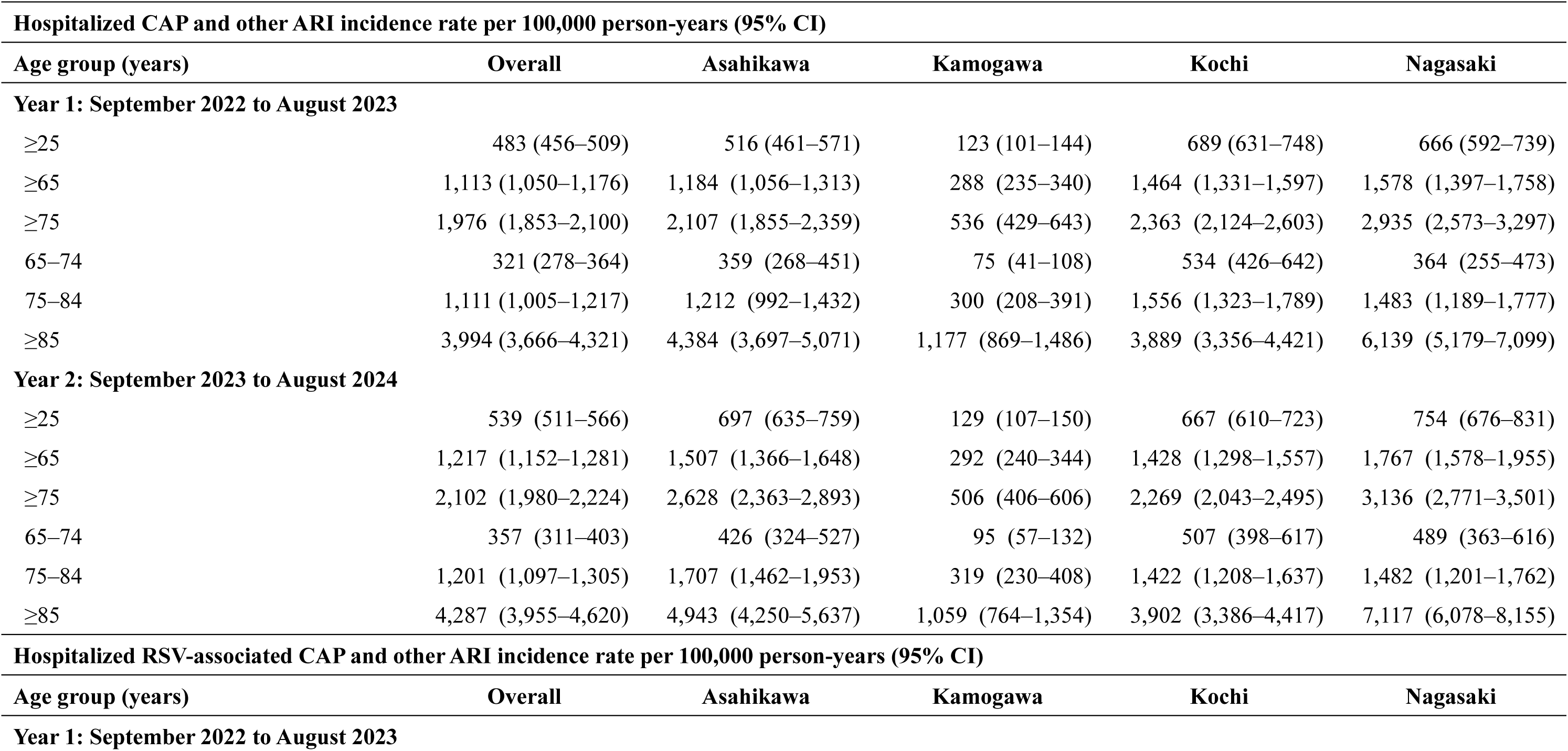
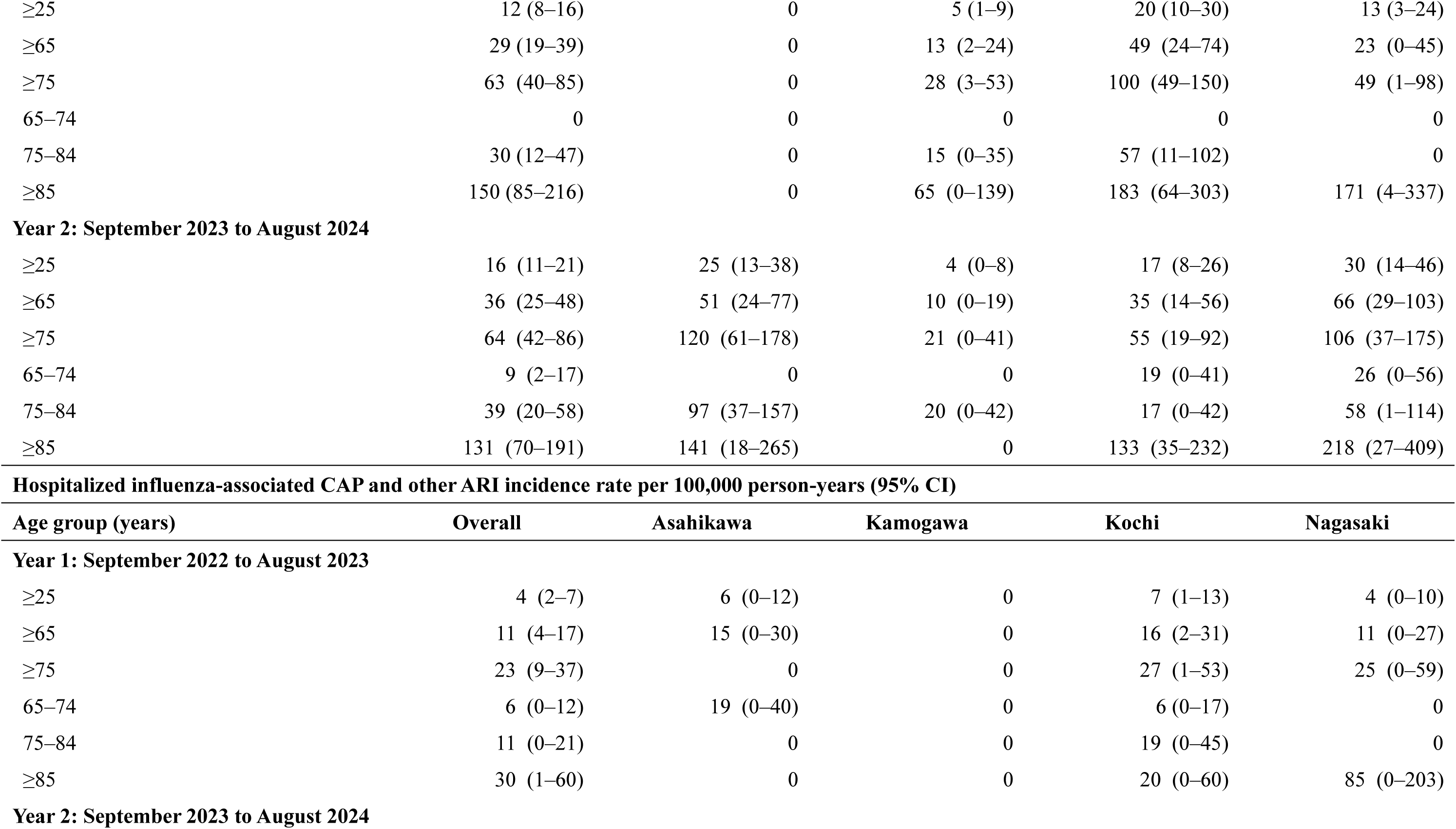
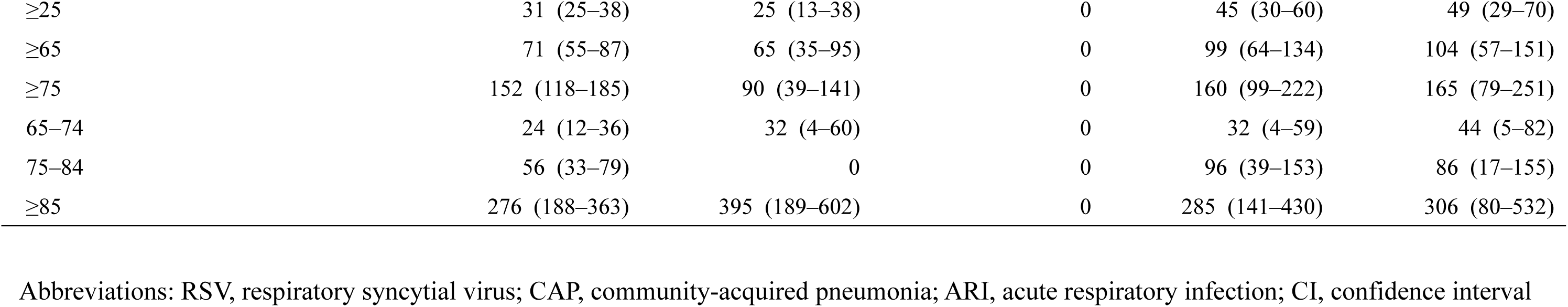
Incidence rates of adults hospitalized with total, RSV-associated, and influenza-associated CAP and other ARI in Japan, September 2022–August 2024.

| **Hospitalized CAP and other ARI incidence rate per 100,000 person-years (95% CI)** |  |  |  |  |  |
| --- | --- | --- | --- | --- | --- |
| **Age group (years)** | **Overall** | **Asahikawa** | **Kamogawa** | **Kochi** | **Nagasaki** |
| **Year 1: September 2022 to August 2023** |  |  |  |  |  |
| ≥25 | 483 (456–509) | 516 (461–571) | 123 (101–144) | 689 (631–748) | 666 (592–739) |
| ≥65 | 1,113 (1,050–1,176) | 1,184 (1,056–1,313) | 288 (235–340) | 1,464 (1,331–1,597) | 1,578 (1,397–1,758) |
| ≥75 | 1,976 (1,853–2,100) | 2,107 (1,855–2,359) | 536 (429–643) | 2,363 (2,124–2,603) | 2,935 (2,573–3,297) |
| 65–74 | 321 (278–364) | 359 (268–451) | 75 (41–108) | 534 (426–642) | 364 (255–473) |
| 75–84 | 1,111 (1,005–1,217) | 1,212 (992–1,432) | 300 (208–391) | 1,556 (1,323–1,789) | 1,483 (1,189–1,777) |
| ≥85 | 3,994 (3,666–4,321) | 4,384 (3,697–5,071) | 1,177 (869–1,486) | 3,889 (3,356–4,421) | 6,139 (5,179–7,099) |
| **Year 2: September 2023 to August 2024** |  |  |  |  |  |
| ≥25 | 539 (511–566) | 697 (635–759) | 129 (107–150) | 667 (610–723) | 754 (676–831) |
| ≥65 | 1,217 (1,152–1,281) | 1,507 (1,366–1,648) | 292 (240–344) | 1,428 (1,298–1,557) | 1,767 (1,578–1,955) |
| ≥75 | 2,102 (1,980–2,224) | 2,628 (2,363–2,893) | 506 (406–606) | 2,269 (2,043–2,495) | 3,136 (2,771–3,501) |
| 65–74 | 357 (311–403) | 426 (324–527) | 95 (57–132) | 507 (398–617) | 489 (363–616) |
| 75–84 | 1,201 (1,097–1,305) | 1,707 (1,462–1,953) | 319 (230–408) | 1,422 (1,208–1,637) | 1,482 (1,201–1,762) |
| ≥85 | 4,287 (3,955–4,620) | 4,943 (4,250–5,637) | 1,059 (764–1,354) | 3,902 (3,386–4,417) | 7,117 (6,078–8,155) |
| **Hospitalized RSV-associated CAP and other ARI incidence rate per 100,000 person-years (95% CI)** |  |  |  |  |  |
| **Age group (years)** | **Overall** | **Asahikawa** | **Kamogawa** | **Kochi** | **Nagasaki** |
| **Year 1: September 2022 to August 2023** |  |  |  |  |  |
| ≥25 | 12 (8–16) | 0 | 5 (1–9) | 20 (10–30) | 13 (3–24) |
| ≥65 | 29 (19–39) | 0 | 13 (2–24) | 49 (24–74) | 23 (0–45) |
| ≥75 | 63 (40–85) | 0 | 28 (3–53) | 100 (49–150) | 49 (1–98) |
| 65–74 | 0 | 0 | 0 | 0 | 0 |
| 75–84 | 30 (12–47) | 0 | 15 (0–35) | 57 (11–102) | 0 |
| ≥85 | 150 (85–216) | 0 | 65 (0–139) | 183 (64–303) | 171 (4–337) |
| **Year 2: September 2023 to August 2024** |  |  |  |  |  |
| ≥25 | 16 (11–21) | 25 (13–38) | 4 (0–8) | 17 (8–26) | 30 (14–46) |
| ≥65 | 36 (25–48) | 51 (24–77) | 10 (0–19) | 35 (14–56) | 66 (29–103) |
| ≥75 | 64 (42–86) | 120 (61–178) | 21 (0–41) | 55 (19–92) | 106 (37–175) |
| 65–74 | 9 (2–17) | 0 | 0 | 19 (0–41) | 26 (0–56) |
| 75–84 | 39 (20–58) | 97 (37–157) | 20 (0–42) | 17 (0–42) | 58 (1–114) |
| ≥85 | 131 (70–191) | 141 (18–265) | 0 | 133 (35–232) | 218 (27–409) |

**Hospitalized influenza-associated CAP and other ARI incidence rate per 100,000 person-years (95% CI)**
| Age group (years) | Overall | Asahikawa | Kamogawa | Kochi | Nagasaki |
| --- | --- | --- | --- | --- | --- |
| **Year 1: September 2022 to August 2023** |  |  |  |  |  |
| ≥25 | 4 (2–7) | 6 (0–12) | 0 | 7 (1–13) | 4 (0–10) |
| ≥65 | 11 (4–17) | 15 (0–30) | 0 | 16 (2–31) | 11 (0–27) |
| ≥75 | 23 (9–37) | 0 | 0 | 27 (1–53) | 25 (0–59) |
| 65–74 | 6 (0–12) | 19 (0–40) | 0 | 6 (0–17) | 0 |
| 75–84 | 11 (0–21) | 0 | 0 | 19 (0–45) | 0 |
| ≥85 | 30 (1–60) | 0 | 0 | 20 (0–60) | 85 (0–203) |
| **Year 2: September 2023 to August 2024** |  |  |  |  |  |
| ≥25 | 31 (25–38) | 25 (13–38) | 0 | 45 (30–60) | 49 (29–70) |
| ≥65 | 71 (55–87) | 65 (35–95) | 0 | 99 (64–134) | 104 (57–151) |
| ≥75 | 152 (118–185) | 90 (39–141) | 0 | 160 (99–222) | 165 (79–251) |
| 65–74 | 24 (12–36) | 32 (4–60) | 0 | 32 (4–59) | 44 (5–82) |
| 75–84 | 56 (33–79) | 0 | 0 | 96 (39–153) | 86 (17–155) |
| ≥85 | 276 (188–363) | 395 (189–602) | 0 | 285 (141–430) | 306 (80–532) |

Abbreviations: RSV, respiratory syncytial virus; CAP, community-acquired pneumonia; ARI, acute respiratory infection; CI, confidence interval

The incidence rates of RSV-associated CAP and other ARI hospitalizations among adults aged ≥65 years was 29 (95% CI: 19–39) and 36 (95% CI: 25–48) per 100,000 person-years. Further age stratification among adults aged ≥65 years revealed an increasing incidence with age, peaking in those aged ≥85 years: 150 (95% CI: 85–216) and 131 (95% CI; 70–191) per 100,000 person-years in the first and second year, respectively.

The incidence rates of influenza-associated CAP and other ARI hospitalizations increased notably from the first to second year. Among adults aged ≥65 years, the incidence rose from 11 (95% CI: 4–17) to 71 (95% CI: 55–87) per 100,000 person-years. The highest rate was observed in those aged ≥85 years: 30 (95% CI: 1–60) and 276 (95% CI: 188–363) per 100,000 person-years in the first and second year, respectively.

Geographically, the incidence rates for total, RSV- and flu-associated CAP and other ARI were generally comparable across Asahikawa, Kochi, and Nagasaki, but consistently lower in Kamogawa. To address this discrepancy, incidence was recalculated, excluding Kamogawa. Among adults aged ≥65, the incidence rates per 100,000 person-years in the first year excluding Kamogawa was 34 (95% CI: 21–48) for RSV-associated, and 15 (95% CI: 6– 24) for influenza-associated CAP and other ARI. In the second year, the corresponding rates were 46 (95% CI: 31–62), and 96 (95% CI: 74–119), respectively.

## 4. Discussion

To our knowledge, this is the first study to evaluate the incidence of RSV-associated hospitalization as well as influenza-associated hospitalizations among adults in Japan during and after the COVID-19 pandemic.

The incidence of adult RSV-associated hospitalizations has been estimated in several countries. However, direct comparisons are challenging due to differences in methodology, patient demographics and hospitalization practices. Among studies using active surveillance during and after the COVID-19 pandemic, our estimates were comparable to those reported from the UK (17), and lower than those from the US and Germany (19, 20). A report from the UK estimated that the incidence of RSV-associated hospitalizations among adults aged ≥65 years ranged from 34 to 54 per 100,000 person-years from the 2017 to 2023 season, and 65 per 100,000 person-years among adults aged ≥75 years during the 2022–2023 season (17), which was comparable to our study results, based on surveillance during the RSV season (week 40 to week 20 of the following year). In contrast, a US study estimated incidence rates of RSV-associated hospitalizations at 98.5 and 160.9 per 100,000 for adults aged ≥65 years during the 2021–2022 and 2022–2023 seasons, respectively (19). A German study reported 134.1 per 100,000 for adults aged ≥65 years from 2021–2023 (20). Both the US and German studies adjusted for test sensitivity (8), and the potential under-detection associated with nasal swab sampling (20, 33).

While abundant epidemiological data on influenza, such as positivity percentage (34, 35) and disease burden (36, 37), are available, data on RSV, including RSV-positivity percentage among hospitalized adults, remain limited in Japan both before and after the COVID-19 pandemic. Two studies conducted in Japan during the post-pandemic era reported RSV-positivity percentages similar to our study (22–24), although none of them reported the incidence rate. One study reported a percentage of 2.1% among hospitalized patients aged ≥50 years with respiratory symptoms (23); another study reported a percentage of 2.6% among out-patients aged ≥60 years with respiratory symptoms from December 2022 to November 2023 (22). Positivity percentages reported prior to the COVID-19 pandemic were slightly higher. The initial APSG-J study conducted from 2011 to 2013 reported an RSV positivity of 5% among inpatients with respiratory symptoms (26). Given that our results may still be influenced by the lingering effects of the COVID-19 pandemic, continued surveillance is essential to accurately assess the burden of RSV in the adult population.

Our study has several limitations. First, recruitment was limited to seven hospitals across four catchment areas, which may limit generalizability. Kamogawa area had lower incidence of hospitalizations with CAP and other ARI, possibly due to strict admission criteria at the hospital, and the rural and relatively isolated location. However, as noted in the Results, this had a limited impact on the overall incidence of RSV-associated hospitalizations and our conclusions. Second, approximately half of the hospitalized CAP and other ARI patients did not undergo multiplex PCR. Nevertheless, the baseline characteristics of those tested and untested were largely compatible (Supplementary Section 2), supporting the generalizability of PCR findings. Third, our estimates of CAP and other ARI hospitalization incidence relied on available governmental data, which may have introduced some uncertainty. Fourth, the incidence of RSV-associated hospitalizations may have been underestimated due to limited sensitivity of our diagnostic testing (19, 20, 33). Fifth, our study included patients with ARI symptoms, possibly missing RSV-associated hospitalizations without overt respiratory symptoms. Some studies estimating incidence included cardiopulmonary disease exacerbation, such as chronic obstructive pulmonary disease (COPD) and congestive heart failure (CHF), regardless of other symptoms (18, 38), while our study included such exacerbations only when accompanied by ARI diagnosis, which may have underestimated the incidence of RSV-associated hospitalizations.

In conclusion, we successfully evaluated the incidence of adult RSV-associated CAP and other ARI hospitalizations in Japan using active surveillance. Our study provides essential evidence to inform discussions on the implementation of novel RSV vaccination policies for older adults.

## Acknowledgement

We thank Rina Shiramizu, Rina Kubo, Yumi Araki, Yukie Kozone, Mao Hamasaki, Megumi Michitsuji and Eriko Yamada from the Institute of Tropical Medicine, Nagasaki University for their technical assistance. We also thank all the laboratory staff and the collaborators at the participating hospitals for their contributions. Adult Pneumonia Study Group—Japan 2 (APSG-J2) are: Yuka Fujita^1^, Toshiaki Fujikane^1^, Tadakatsu Tsuji^1^, Yasuhiro Yamazaki^1^, Kazushi Doshita^1^, Satoshi Endo^1^, Yoshitsugu Narumi^1^, Keiichi Nakamura^1^, Hiraku Yanada^1^, Toshiyuki Tenma^1^, Taeka Naraoka^1^, Ko Takahashi^1^, Minami Kaneko^1^, Masashi Ueda^1, 2^, Yuji Akiba^2^, Yutaka Nishigaki^2^, Yoshihiro Kazebayashi^2^, Katsuya Tsujie^2^, Kiichi Nitanai^2^, Akari Yagita^2^, Maya Ikeda^2^, Kei Nakashima^3^, Hiroyuki Ito^3, 20^, Akiyuki Sato^4^, Yosuke Ebisu^5^, Masayuki Nogi^4^, Akihito Yoshida^4^, Naoto Hosokawa^5^, Ryosuke Osawa^5^, Yoshiro Hayashi^6^, Toshiyuki Karumai^6^, Kohei Takimoto^6^, Atsushi Shiraishi^7^, Yoshihito Otsuka^8^, Tomohisa Watari^8^, Masayuki Ishida^9^, Hiroshi Nakaoka^9^, Eiji Takeuchi^10^, Nobuo Hatakeyama^10^, Hisanori Machida^10^, Yoshio Okano^10^, Naoki Kadota^10^, Michihiro Kunishige^10^, Seiya Ichihara^10^, Yugo Matsumura^10^, Hiroki Takahashi^10^, Kaori Nii^10^, Norichika Asoh^11^, Yoshiko Tsuchihashi^11^, Kei Matsuki^11^, Taro Kikuchi^11^, Toyomitsu Sawai^12^, Daichi Noritomi^12^, Yusei Fukushima^12^, Seiya Kaneko^12^, Koichi Kawasaki^12^, Shun Morimitsu^12^, Hiroshi Gyotoku^12^, Yudai Inadomi^12^, Koichi Hayakawa^13^, Kensuke Takahasi^13^, Yoshihiro Aoki^13^, Masayoshi Takeno^14^, Shinobu Osanai^15^, Ataru Igarashi^16^, Eileen M Dunne^17^, Claudia Schwarz^17^, Bradford D. Gessner^17^, Elizabeth Begier^17^, Shuhei Ito^18^, Koya Ariyoshi^19^, Konosuke Morimoto^20^, Haruka Maeda^20^, Bhim Gopal Dhoubhadel^20^, Shingo Masuda^19, 20^.

1. Department of Respiratory Medicine, National Hospital Organization Asahikawa Medical Center, Asahikawa, Japan
2. Department of Respiratory Medicine, Asahikawa-Kosei General Hospital, Asahikawa, Japan
3. Department of Pulmonology, Kameda Medical Center, Kamogawa, Japan.
4. Department of General Internal Medicine, Kameda Medical Center, Kamogawa, Japan.
5. Department of Infectious Diseases, Kameda Medical Center, Kamogawa, Japan.
6. Department of Intensive Care Medicine, Kameda Medical Center, Kamogawa, Japan.
7. Emergency and Trauma Center, Kameda Medical Center, Kamogawa, Japan
8. Department of Clinical Laboratory, Kameda Medical Center, Kamogawa, Japan
9. Department of Infectious Disease Medicine, Department of Respiratory Medicine, Chikamori Hospital, Kochi, Japan
10. Department of Respiratory Medicine, National Hospital Organization Kochi Hospital, Kochi, Japan
11. Department of Internal Medicine, Juzenkai Hospital, Nagasaki, Japan
12. Department of Respiratory Medicine, Nagasaki Harbor Medical Center, Nagasaki, Japan
13. Department of Emergency Medicine, Nagasaki Harbor Medical Center, Nagasaki, Japan
14. Department of Cardiovascular Medicine, Nagasaki Harbor Medical Center, Nagasaki, Japan
15. Division of Respiratory Medicine and Neurology, Department of Internal Medicine, Asahikawa Medical University, Asahikawa, Japan
16. Department of Health Policy and Public Health, Graduate School of Pharmaceutical Sciences, The University of Tokyo, Tokyo, Japan
17. Pfizer Vaccines, Collegeville, PA, USA
18. Vaccine medical Affairs, Pfizer Japan inc., Tokyo, Japan
19. Department of Clinical Medicine, Institute of Tropical Medicine, Nagasaki University, Nagasaki, Japan
20. Department of Respiratory Infections, Institute of Tropical Medicine, Nagasaki University, Nagasaki, Japan

## Data availability

The data supporting the findings of this study are not available as consent for data sharing was not obtained.

## Author contributions

KM, KA, and BG conceived and designed the study. YF, YA, YN, KN, HI, MN, YO, MI, ET, NA, TS, and KH collected the data. HM analyzed the data, and HM, KM, KA, EMD, CS, BG, EB, and SI interpreted the results. HM, KM and KA drafted the manuscript, and all authors critically revised the manuscript and approved the final version.

## Ethics

This study was approved by the Institutional Review Board of the Institute of Tropical Medicine, Nagasaki University with a centralized review (Approval No.: 220616247). With this approval, the investigators at the six sites obtained permission to conduct the study. In the remaining site, the study was approved by its individual review board following the approval by the centralized review (Approval No. :22-07-F).

