## Supplementary Material for "Estimating Incidence of Respiratory Syncytial Virus- and Influenza Virus-Associated Hospitalizations with Community-Acquired Pneumonia and Other Acute Respiratory Infection Among Adults in Japan During and After the COVID-19 Era: A Multicenter Active Surveillance Study (APSG-J2)"

### Supplementary Materials

**Supplementary Section S1: Inclusion and exclusion criteria for APSG-J2**

Inclusion criteria were as follows: patients aged ≥18 years who met at least one of the following conditions: (1) clinical signs or symptoms of acute respiratory infection (ARI), (2) a diagnosis of pneumonia or ARI made by the attending physician, or (3) an exacerbation of underlying cardiopulmonary disease with a concurrent ARI diagnosis. Radiographic evaluation (chest X-ray or computed tomography [CT]) was also required. ARI signs or symptoms included any of the following: fever ≥38°C, cough, sputum production, pleuritic chest pain, dyspnea, tachypnea (respiratory rate >20/min), hypoxemia ($SpO_2$ <93%), nasal congestion, rhinorrhea, sore throat, and hoarseness.
Exclusion criteria included: a positive severe acute respiratory syndrome coronavirus 2 (SARS-CoV-2) test result at admission; pneumonia more than 48 hours after hospital admission; residence outside the hospital's designated catchment area; prior enrollment in the study within the past 14 days; and diagnosis of pulmonary tuberculosis or chronic pulmonary nontuberculous mycobacterial infection. At the Kamogawa site, patients from outside the catchment area were included to calculate the proportion of RSV- or influenza-associated CAP/ARI hospitalizations among CAP/ARI patients who underwent multiplex PCR due to their high numbers but were excluded from incidence estimates.

**Supplementary Section S2: Characteristics of adults hospitalized with CAP and other ARI, with and without multiplex PCR, and those with only age, sex, and diagnosis information available, in Japan, September 2022–August 2024**

Supplementary Table 1 shows the characteristics of adults hospitalized with community-acquired pneumonia (CAP) and other acute respiratory infection (ARI), comparing those who underwent multiplex polymerase chain reaction (PCR) with those who did not. Overall, patients who received multiplex PCR were generally similar to those who were not tested. However, untested patients were slightly older (aged ≥65 years: 90.6% vs. 87.7%), more likely to be female (40.5% vs. 35.9%), less likely to reside in a nursing home (17.4% vs. 30.1%), and less likely to have one or more underlying medical conditions (77.2% vs. 82.2%) (Supplementary Table 1).

**Table S1. Characteristics of adults hospitalized with CAP and other ARI, with and without multiplex PCR, and those with only age, sex, and diagnosis information available, in Japan, September 2022–August 2024**

| | CAP and other ARI hospitalizations with multiplex PCR, no. (%) | CAP and other ARI hospitalizations without multiplex PCR, no. (%) | CAP and other ARI hospitalizations of patients in the Declined Consent Arm[a], no. (%) |
|---|---|---|---|
| Overall | 1,502 (100) | 1,366 (100) | 179 (100) |
| Median age (IQR), years | 81 (73–88) | 82 (75–89) | 80 (74–86) |
| Age group in years | | | |
| 18–64 | 184 (12.3) | 129 (9.4) | 17 (9.5) |
| ≥65 | 1,318 (87.7) | 1,237 (90.6) | 162 (90.5) |
| Sex | | | |
| Men | 963 (64.1) | 813 (59.5) | 122 (68.2) |
| Women | 539 (35.9) | 553 (40.5) | 57 (31.8) |
| Underlying medical conditions[b] | | | |
| Any | 1,235 (82.2) | 1,055 (77.2) | N/A |
| Asplenia | 1 (0.1) | 0 | N/A |
| Cancer | 234 (15.6) | 245 (17.9) | N/A |
| Leukemia | 2 (0.1) | 3 (0.2) | N/A |
| Lymphoma | 10 (0.7) | 8 (0.6) | N/A |
| Multiple myeloma | 1 (0.1) | 6 (0.4) | N/A |
| Immunosuppressive drug use | 74 (4.9) | 68 (5.0) | N/A |
| Organ transplantation | 2 (0.1) | 0 | N/A |
| Diabetes mellitus | 348 (23.2) | 317 (23.2) | N/A |
| Nephrotic syndrome | 3 (0.2) | 5 (0.4) | N/A |
| Chronic heart failure | 264 (17.6) | 266 (19.5) | N/A |
| Chronic heart disease other than chronic heart failure | 343 (22.8) | 284 (20.8) | N/A |
| Chronic lung disease (including asthma) | 604 (40.2) | 353 (25.8) | N/A |
| Chronic renal failure | 146 (9.7) | 114 (8.3) | N/A |
| Chronic live disease | 68 (4.5) | 55 (4.0) | N/A |
| Obesity (BMI ≥40) | 4 (0.3) | 1 (0.1) | N/A |
| Cerebrospinal fluid leakage | 1 (0.1) | 1 (0.1) | N/A |

| | | | |
|---|---|---|---|
| Cerebrovascular disease | 271 (18.0) | 261 (19.1) | N/A |
| Current smokers | | | |
| Yes | 134 (8.9) | 145 (10.6) | N/A |
| No | 1,287 (85.7) | 976 (71.4) | N/A |
| Unknown | 81 (5.4) | 245 (17.9) | N/A |
| Nursing-home residents | 262 (17.4) | 411 (30.1) | N/A |
| Nursing care level[c] | | | |
| Independent | 807 (53.7) | 595 (43.6) | N/A |
| Support (1–2) | 145 (9.7) | 99 (7.2) | N/A |
| Care (1–5) | 548 (36.5) | 654 (47.9) | N/A |
| Unknown | 2 (0.1) | 18 (1.3) | |
| Prior hospitalization within the past 90 days | | | |
| Yes | 290 (19.3) | 258 (18.9) | N/A |
| No | 1,188 (79.1) | 1,056 (77.3) | N/A |
| Unknown | 24 (1.6) | 52 (3.8) | N/A |
| Preceding antibiotics use within 14 days | | | |
| Yes | 298 (19.8) | 229 (16.8) | N/A |
| No | 1,190 (79.2) | 1,100 (80.5) | N/A |
| Unknown | 14 (0.9) | 37 (2.7) | N/A |
| Study period | | | |
| Year 1 (September 2022, to August 2023) | 754 (50.2) | 595 (43.6) | N/A |
| Year 2 (September 2023, to August 2024) | 748 (49.8) | 771 (56.4) | N/A |
| Area | | | N/A |
| Asahikawa | 218 (14.5) | 572 (41.9) | N/A |
| Kamogawa | 198 (13.2) | 274 (20.1) | N/A |
| Kochi | 782 (52.1) | 180 (13.2) | N/A |
| Nagasaki | 304 (20.2) | 340 (24.9) | N/A |
| Diagnosis | | | |
| Community-acquired pneumonia | 1,361 (90.6) | 1,235 (90.4) | 152 (84.9) |
| Other acute respiratory infection | 141 (9.4) | 131 (9.6) | 27 (15.1) |

Abbreviations: CAP, community-acquired pneumonia; ARI, acute respiratory infection; PCR, polymerase chain reaction; IQR, interquartile range; BMI, body mass index.

a: For patients who declined to participate were included in the Declined Consent Arm, only age, sex, and diagnosis information of them were registered (See Supplementary Figure 1).
b: No patients had sickle cell disease, HIV infection, Hodgkin's disease, or cochlear implant.

c: Nursing care level was determined based on the Long-Term Care Insurance System by the Ministry of Health, Labour and Welfare. Details are provided in Supplementary Table 3.

**Table S2. Sample types for multiplex PCR from adult hospitalizations with community-acquired pneumonia and other acute respiratory infections in Japan, September 2022–August 2024**

| Sample type, no. | Overall, no. (%) | Sep 2022–Aug 2023, no. (%) | Sep 2023–Aug 2024, no. (%) |
|---|---|---|---|
| Any | 1,502 | 754 | 748 |
| Sputum | 1,328 (88.4) | 698 (92.6) | 630 (84.2) |
| Saliva | 386 (25.7) | 89 (11.8) | 297 (39.7) |
| Nasopharyngeal swab | 108 (7.2) | 23 (3.1) | 85 (11.4) |

Abbreviations: PCR, polymerase chain reaction.

**Table S3. Classification of Nursing Care Level in Japan**

| Care level | |
|---|---|
| Independent | Capable of performing daily activities without assistance. |
| Support 1 | Capable of performing daily activities independently but requires occasional assistance. |
| Support 2 | Requires some assistance with daily activities, with a high potential for improvement without the need for nursing care. |
| Nursing Care 1 | Experiences unsteadiness when standing or walking.<br>Requires partial assistance with daily activities such as toileting and bathing. |
| Nursing Care 2 | Has difficulty standing or walking independently.<br>Requires partial to full assistance with daily activities such as toileting and bathing. |
| Nursing Care 3 | Unable to stand or walk.<br>Requires full assistance with daily activities including toileting, bathing, and dressing/undressing. |
| Nursing Care 4 | Requires full assistance in all aspects of daily life, including toileting, bathing, and dressing/undressing. |
| Nursing Care 5 | Requires full assistance in all aspects of daily life and has significant difficulty with communication. |

This classification is defined by the Ministry of Health, Labour and Welfare in Japan.

**Figure S1. Study flow and surveillance arm**

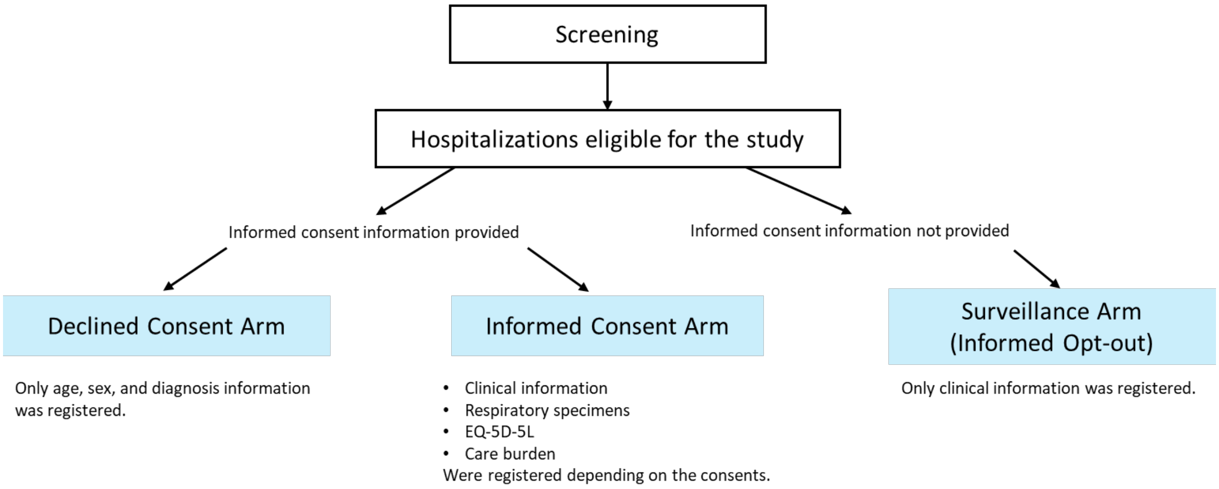

Following screening, hospitalizations eligible for the study were categorized into three groups based on the informed consent procedures. Patients who explicitly declined participation were assigned to the Declined Consent Arm. Those who provided informed consent were included in the Informed Consent Arm. Patients who were not directly approached for consent, in accordance with the Japanese guidelines on informed opt-out, were assigned to the Surveillance Arm (Informed Opt-out).
